# Plasma gangliosides correlate with disease stages and symptom severity in Huntington’s disease carriers

**DOI:** 10.1101/2025.03.21.25324430

**Authors:** D. Galleguillos, Y. Zhao, B. Pan, B. Vandermeer, A. Zaidi, YN Al Hamarneh, J. Sarna, O. Suchowersky, J. Curtis, S. Sipione

## Abstract

**Background:** Gangliosides - glycosphingolipids that modulate cell signaling and neuronal functions - are decreased in Huntington’s disease (HD) models and patients’ brains. Restoring ganglioside GM1 has therapeutic benefits in HD mice, slowing neurodegeneration and improving symptoms. This suggests gangliosides might contribute to HD pathogenesis. However, their link to disease severity and progression in patients remains unclear.

**Objectives:** This study examined plasma ganglioside differences between HD gene carriers and controls, and their prognostic potential.

**Methods:** Plasma gangliosides were quantified in 67 HD carriers and 46 healthy participants, using liquid chromatography-tandem mass spectrometry. Statistical modelling assessed associations with clinical measures and prognostic potential.

**Results:** Levels of most gangliosides were similar between groups, but GM3 was higher and GT1b lower in HD carriers. Within the HD group, higher GM2 levels correlated with better cognition, and higher GM1 and GD1a with greater functional capacity and independence. Higher GM1 predicted HD status, but its decline and an increase in GD3 were strongly associated with disease progression. Individual gangliosides had limited disease classification ability.

**Conclusions:** The correlation between higher GM2, GD1a and GM1 and milder symptoms suggests a protective role of these gangliosides in HD. The association between higher GM1 levels and HD status, along with its decline predicting disease progression, suggests GM1 increase may be a compensatory neuroprotective mechanism that deteriorates over time. While plasma gangliosides are not strong disease classifiers, our findings provide novel insights into their role in HD progression and prognostic potential.

## INTRODUCTION

Gangliosides - sialic acid-containing glycosphingolipids – are crucial for neuronal function, signal transduction and cell-cell interactions, and have gained attention in the context of neurodegenerative diseases due to their general neuroprotective properties (1). The importance of gangliosides for brain health is underscored by the fact that loss of function mutations in enzymes of the ganglioside biosynthetic pathway lead to severe early onset neurodegenerative disorders, including the early onset symptomatic epileptic syndrome (2) and a complex form of hereditary spastic paraplegia (3, 4). Alterations in brain ganglioside composition, including decreases in specific gangliosides, have been reported in aging (5-8) and in common neurodegenerative conditions such as Alzheimer’s disease, Parkinson’s disease, amyotrophic lateral sclerosis and Huntington’s disease (9-13). These findings raise the question of whether a dysregulation of ganglioside metabolism contributes to the onset, pathogenesis or progression of these diseases.

Several lines of evidence suggest that gangliosides play a role in Huntington’s disease (HD), a dominantly inherited neurodegenerative disorder caused by the misfolding and aggregation of mutant huntingtin (mHTT). First, GM1 and other major brain gangliosides are reduced in post-mortem HD brains (11), as well as in fibroblasts isolated from HD patients and in HD cell and mouse models (10). Second, in striatal cells derived from HD knock-in mice, a ∼25% decrease in GM1 levels compared to control cells is sufficient to increase cell susceptibility to apoptosis (10). Third, replenishing gangliosides via GM1 administration promotes cell survival *in vitro* (10) and exerts profound disease-modifying and therapeutic effects in HD animal models, leading to reduced neurodegeneration and dramatic improvements in motor and non-motor dysfunctions (14, 15). These studies highlight the neuroprotective role of endogenous gangliosides in HD models, suggesting that ganglioside depletion may contribute to disease progression, and point to GM1 administration as a potential therapeutic strategy.

Despite this compelling preclinical evidence, it remains unclear whether ganglioside levels in human HD gene carriers correlate with disease onset or progression. Moreover, their potential as biomarkers for HD status or disease trajectory has yet to be determined.

This study aimed to: 1) determine whether plasma ganglioside levels differ between healthy controls and HD gene carriers at different disease stages, and 2) assess correlations between plasma gangliosides and clinical measures of disease progression.

To address these questions, we developed a high-performance liquid chromatography tandem mass spectrometry (HPLC-MS/MS) method to quantify plasma gangliosides in a cohort of 67 HD carriers and 46 age-matched healthy controls. Using regression models and machine learning approaches, we assessed the relationship between different gangliosides and HD status and progression. Our data suggest that plasma gangliosides have prognostic value and provide novel insights into their potential role in HD pathophysiology and disease progression.

## METHODS

### Study design

This study assessed plasma ganglioside changes in HD and their relevance as biomarkers of HD status and severity. A total of 113 volunteers were recruited across three clinical centers at the University of Alberta (Edmonton, Canada), the University of Calgary (Canada) and the University of Montreal (Canada). Exclusion criteria for all participants included current or past history (within the last 5 years) of autoimmune disorders, malignancy, use of steroids, immune-suppressive medications and/or statins, as well as any psychiatric disorder. On the day of the visit, a battery of neurological tests was conducted in all HD gene carriers and their scores on the Unified Huntington Disease Rating Scale (UHDRS), including Total Motor Score (TMS), Total Functional Capacity (TFC) and Independence Scale (IS), as well as Montreal Cognitive Assessment (MoCA) scores were collected. CAP scores were calculated as follows: (age at entry) x (CAG – 34). None of the volunteers had significant cognitive impairment (MMSE ≥24). HD-carriers were grouped into early pre-manifest (>8 years to onset, TMS = 0-3, TFC = 13, CAP < 368), late pre-manifest (≤ 8 years to onset, TMS ≤ 6, TFC = 13, CAP ≥ 368), mild symptomatic HD (TFC = 10-13) and moderate symptomatic HD (TFC < 10), in line with TRACK-HD (16, 17). Ethical approval was obtained from the University of Alberta Health Research Ethics Board (Pro00067917). All data were de-identified and assigned an alpha-numerical code. Blood samples were collected as specified below with written informed consent of the participants.

### Chemicals and reagents

Bovine brain monosialoganglioside-GM3 (GM3), trisialoganglioside-GD3, disialoganglioside-GD1b, and trisialoganglioside-GT1b were obtained from Axxora, LLC (Famingdale, NY 11735, USA). Ganglioside GM2 was purchased from Adipogen (San Diego, CA 92121-4721, USA). Monosialoganglioside-GM1 (GM1) from porcine brain was obtained from TRB Chemedica Int., and disialoganglioside-GD1a (GD1a) from bovine brain was purchased from Enzo Life Sciences. The internal standards N-*omega*-CD_3_-octadecanoyl monosialoganglioside GM_1_ (NH_4_^+^salt, GM1-d_3_) and N-omega-CD_3_-octadecanoyl monosialoganglioside GM_3_ (NH_4_^+^salt, GM3-d_3_) were supplied by Matreya LLC (State College, PA, USA). Ammonium acetate (99%), HPLC grade methanol, Optima LC/MS grade water and LC/MS grade acetonitrile were purchased from the Fisher Scientific Company (Ottawa, Canada).

### Blood sample collection and plasma separation

Blood samples were collected the same day of the visit. Study participants were instructed to abstain from consuming dairy products in the 12 h prior to their visit, but fasting was not required. Up to 24 ml of venous blood was collected with standard phlebotomy techniques into 8.5 ml acid citrate dextrose (ACD) vacutainer blood collection tubes (Becton Dickinson, #364606). Samples collected at the University of Alberta were immediately transported to the Canadian Biosample Repository (Edmonton, AB, Canada) where plasma was separated by blood centrifugation at 500 x g for 10 min, aliquoted in cryovials and stored at -80°C until ganglioside analysis. Blood samples collected at the University of Calgary and at the CHUM Research Centre were shipped on the same day in insulated packages (Saf-T-Pak STP-309) and processed within 24-30 h.

### Extraction of gangliosides from plasma

Plasma samples were thawed on ice water before use. 150 µl of plasma were spiked with 150µl of an internal standard mixture of GM3-d3 and GM1-d3 (1µg/ml and 0.5µg/ml, respectively). One ml of methanol was then added to each sample, the mixture was vortexed for 1 min and then centrifuged at 10,000 rpm for 15 min. The resulting supernatant was collected, and the pellet was re-extracted once with 400µl of methanol, vortexed, and centrifuged as before. The combined supernatant was dried under a stream of nitrogen and re-dissolved into 150 µl of 1/1 (*v/v*) methanol/acetonitrile for further analysis by LC/MS-MS. All plasma samples were extracted in triplicates.

### HILIC LC/MS-MS analysis

Standards and sample extracts were analyzed using an Agilent 1200 series system coupled to a 3200 QTRAP mass spectrometer (AB Sciex, Canada). An Xbridge HILIC column (150 x 2.1mm, 3.5 µm, Waters) was used for liquid chromatography (LC) separation. The mobile phase A was 1:1 acetonitrile/50mM aqueous ammonium acetate, and mobile phase B was acetonitrile. The gradient was as follows: 0.1min 5% A, increased to 80% A over 20 min, hold at 80% A for 2 min, then back to 5% A at 22.1 min for 8 min to re-equilibrate the column. The injection volume was 10 µl and the flow rate was 300 µl/min. The overall cycle time was 30 min/injection. A valve was programmed by the software to divert the LC effluent to waste before and after the selected retention time window from 8 to 22 min. For mass spectrometry analysis, a TurboIonSpray® source was used in negative ion mode with nitrogen as the curtain gas, drying gas and nebulizing gas. The ion spray voltage was -4500V and the ion source temperature was 400°C. Multiple reaction monitoring (MRM) mode was used for all the investigated compounds. Analyst 1.4.2 software was used to acquire and process data. A six-point matrix-adjusted calibration curve was established for measuring gangliosides in plasma. To achieve this, equal amounts of plasma from each sample were pooled together. The pooled plasma samples were extracted using the previously described procedure. The combined extract solutions were spiked with calibration standards at five different concentrations. A calibration curve was plotted by the concentration of each standard ganglioside spiked versus the peak area ratio of each ganglioside to its respective internal standard. GM3-d3 (transition ion pair 1182.8/290) was used as the internal standard for GM3, and GM1-d3 (MRM transition ion pair 1547.9/290) was used as the internal standard for all other gangliosides. For individual GM3 and GD3 species quantification, the peak area ratio of the analyte to the internal standard was plotted instead of concentration, due to the unavailability of individual species standards. To determine the extraction recovery, 150 µl of pooled plasma samples were spiked with a mixture of gangliosides GM3, GM2, GM1, GD3, GD1a, GD1b, and GT1b. Analytes were spiked at both low and high concentration levels prior to extraction. The spiked plasma samples were then extracted using the same procedure. The recovery was calculated as:

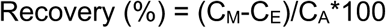

Where: C_M_ the measured concentration of spiked samples; C_E_ the concentration of endogenous ganglioside; C_A_ the concentration of added ganglioside.

### Statistical analysis

All analysis were performed using R 3.4.0 (Vienna, Austria; https://www.R-project.org/) and SAS 9.4 software (SAS Institute Inc. Cary, NC, USA). Patients’ demographic information and clinical characteristics were summarized using descriptive statistics. Categorical variables were reported as frequency (percentage), while continuous variables were summarized using mean ± standard deviation (SD).

Continuous variables were analyzed using *t*-test for normally distributed data, or the Wilcoxon rank-sum test when appropriate. Clinical measurements including MoCA, TMS, TFC, IS, CAPE and CAG repeat size were analyzed using multivariate analysis of variance (MANOVA) to test for significant differences between groups. Correlations between plasma ganglioside levels and clinical measures were assessed using Pearson’s (for normal data) and Spearman’s (for skewed data) coefficients. 95% confidence intervals (CI) were estimated either based on exact distribution or using the empirical likelihood method, as appropriate. A univariate linear mixed-effect model was used to quantify ganglioside impact on clinical measures. This model accounted for participant-specific variability as a random effect, reporting regression coefficients with standard error (S.E), 95% confidence intervals and *p*-values. Univariate logistic regression identified potential ganglioside biomarkers, followed by multivariate logistic regression to determine independent predictors of disease classification and progression. Variables included in the multivariate model were selected based on results from univariate level analysis and Sure Screening procedure (18-20). Receiver Operating Characteristic (ROC) analysis was conducted to assess the overall diagnostic ability of gangliosides. *P*-values were adjusted for multiple comparisons by Bonferroni correction (threshold of significance: 0.05/number of comparisons). Assumptions for all statistical models were assessed and verified prior to analysis.

### Data sharing

Original data are available upon request.

## RESULTS

### Characteristics of study participants

A total of 113 volunteers (54 males and 59 females) was recruited across three clinical sites (Edmonton, Calgary and Montreal). Sixty-seven (46.3% females) carried an expanded *HTT* allele (HD gene carriers). Forty-six age-matched healthy controls (60.8% females) were recruited from unaffected family members and other volunteers. Group demographics are shown in Table 1. HD carriers included pre-manifest (early and late) and symptomatic individuals, the latter been divided into two groups, mild and moderate symptomatic, based on their TFC score. HD carriers’ CAG repeat expansions ranged from 38 to 52. The mean age at visit was 49.2 years for HD-carriers and 52.5 for controls. Disease severity in HD carriers was scored using the Unified Huntington’s Disease Rating Scale (UHDRS), Total Motor Score (TMS), Total Functional Capacity (TFC) and Independence Score (IS) (21). Disease burden (CAG-AGE-Product, CAP) was calculated according to Penney et al. (22). All HD carriers also received a Montreal Cognitive Assessment (MoCA) (23). Clinical symptoms were progressively more severe across different HD groups, as shown by MoCA scores (range 12.0 –30.0, *p*<0.0001), TMS (range 0.0 – 65.0, *p*<0.0001), TFC (range 3.0 – 14.0, *p*<0.0001), IS scores (range 55.0 – 100.0, *p*<0.0001) and CAP (range 156.0 – 702.0, *p*<0.0001) (**Table 1**).

**Table 1.**
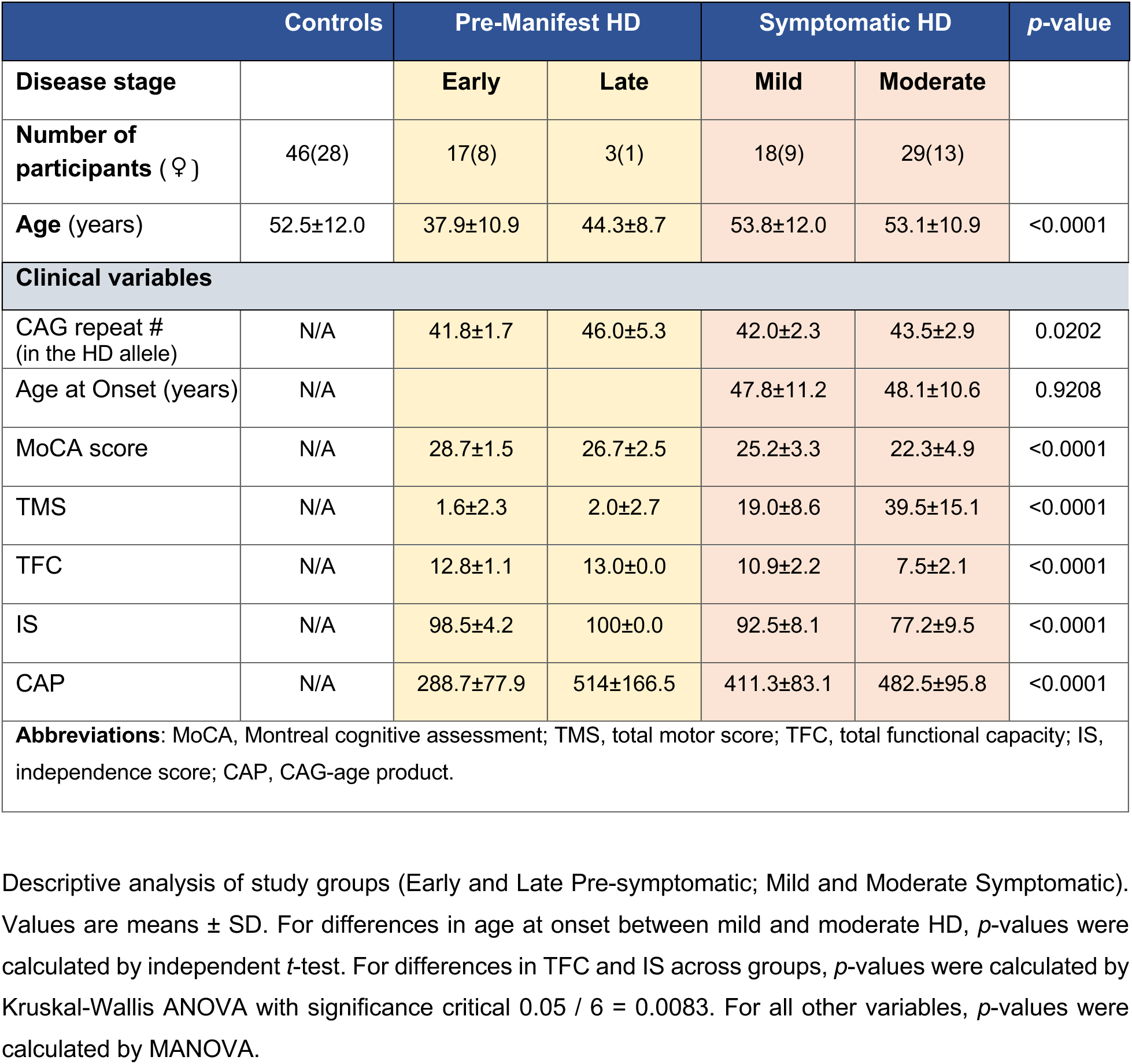
Study participants’ demographic and clinical characterization.

Blood samples collected at the three different clinical locations were processed to separate plasma for ganglioside analysis within 24-30 h from collection.

### HILIC LC/MS-MS separation and analysis provide an accurate measurement of major ganglioside classes in human plasma

The major ganglioside classes present in plasma and brain are shown in **Fig. 1A**. They share a common biosynthetic pathway and differ in the composition of their oligosaccharide headgroups, which determine their structural and functional properties.

**Figure 1.**
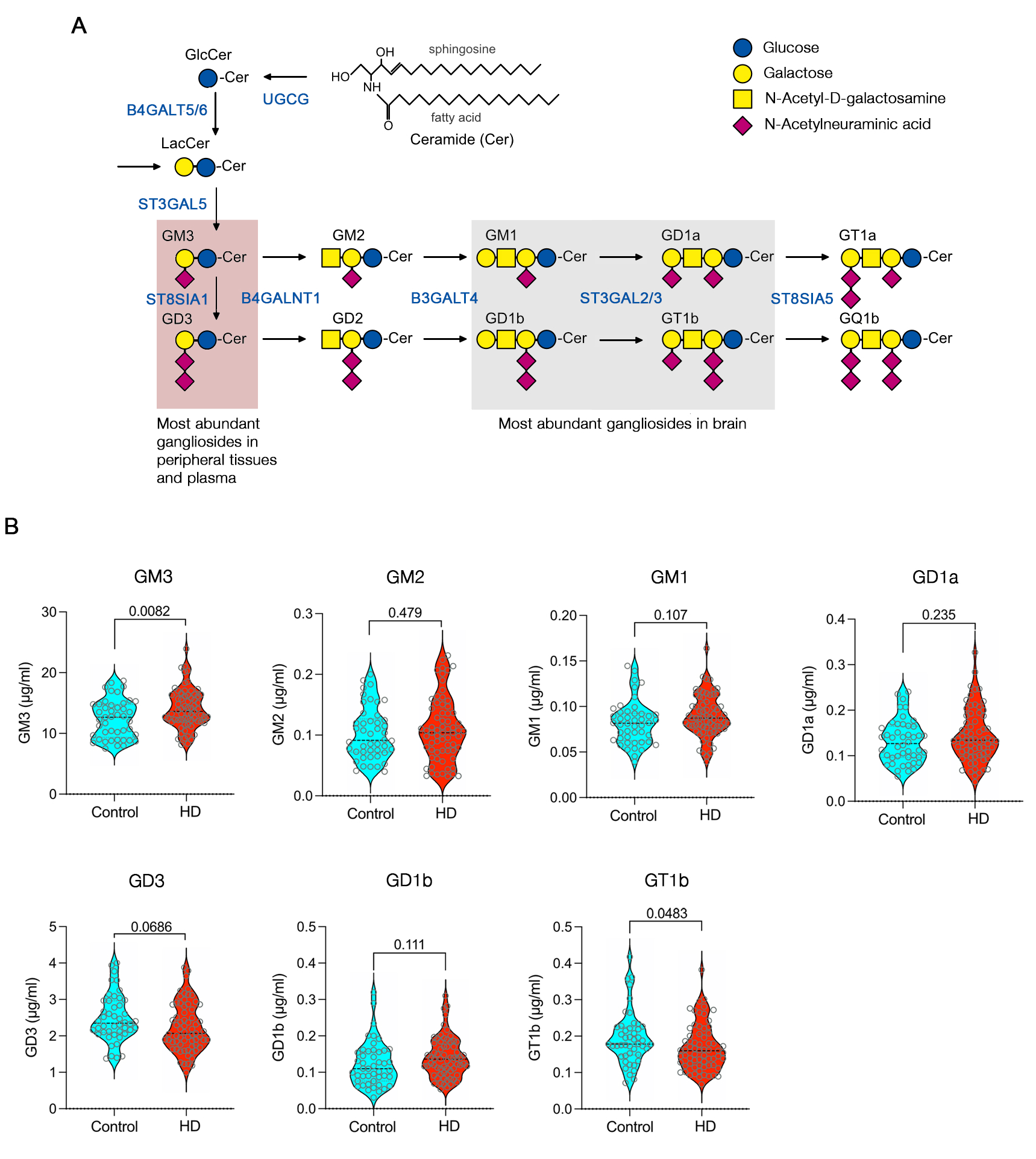
Plasma ganglioside levels in control and HD volunteers. A. Schematic representation of the ganglioside biosynthetic pathway and related enzymes. B. Violin plots displaying median values and the distribution of ganglioside concentrations in control and HD carrier groups. *p-*values from univariate logistic regression are shown above each plot.

The HILIC/MS/MS method was optimized for sample extraction method, HPLC column selection, mobile phase composition, injection solvent, and for MS parameters. Liquid-liquid extraction was tested using various organic solvents, including methanol, acetonitrile, isopropanol, water, chloroform/methanol (1/2), and combinations with and without formic acid. Methanol gave the highest extraction efficacy, which was consistent with literature (24). Multiple HILIC columns, mobile phase compositions and pH were tested. A WatersXbridge HILIC column using a gradient of acetonitrile and acetonitrile/water with 50mM ammonium acetate (50/50) as described above enabled us to separate GM3, GM2, GM1, GD3, GD1a, GD1b, and GT1b, except for GM3/GD1 which were partially separated. **Table S1** lists all MS-related optimized parameters. After evaluating the matrix effect (**Table S2**), a matrix-adjusted calibration curve was used for quantification of gangliosides in plasma. The well-developed HILIC/MS/MS method was assessed by a recovery test (**Table S3**). Recoveries ranged from 92%% to 114% for all gangliosides spiked at high and low concentrations with only one exception (GD1a low concentration spike, 76% recovery).

### Plasma ganglioside levels in HD carriers versus controls

We quantified the abundance of gangliosides GM3, GM2, GM1, GD3, GD1a, GD1b and GT1b in the plasma of control and HD gene carriers. GM3 was the most abundant plasma ganglioside, followed by GD3, confirming previous reports (25, 26). GM2 GM1, GD1a, GD1b and GT1b concentrations were 2 orders of magnitude lower (**Fig. 1B**). No associations were found between plasma gangliosides levels and age (supplementary **Fig. S1**) or biological sex (supplementary **Fig. S2**), therefore data from males and females were pooled together. In the HD-carrier group, no association was found between plasma ganglioside levels and CAG repeat length in the mutant allele (supplementary **Fig. S3**).

Ganglioside concentrations varied within a relatively wide range in both healthy controls and HD-carriers (**Fig. 1B**). Compared to controls, HD gene carriers had higher GM3 (*p* = 0.0082; CI 0.0493 – 0.3054), and lower GT1b levels (*p* = 0.0483; CI -11.7209 - -0.1864). No differences were observed between the two groups for all other gangliosides.

### Higher GM2, GM1 and GD1a levels correlate with milder symptoms

Studies in HD cell and animal models have shown that increasing GM1 levels results in neuroprotection and a dramatic improvement of disease symptoms (10, 14, 15). Therefore, we investigated whether plasma levels of gangliosides in HD gene carriers might correlate with age at HD onset, disease burden (CAP score) or symptoms severity. We did not find significant correlations with age at onset and CAP scores (**Fig. 2** and supplementary **Table S4**). However, higher GM2 levels correlated with higher MoCA scores (r = 0.3045, *p* = 0.0122); higher GM1 levels correlated with greater total functional capacity (TFC: r = 0.2836, *p* = 0.0201) and independence (IS: r = 0.3460, *p* = 0.0041); and GD1a levels correlated with IS (r = 0.2742, *p* = 0.0248).

**Figure 2.**
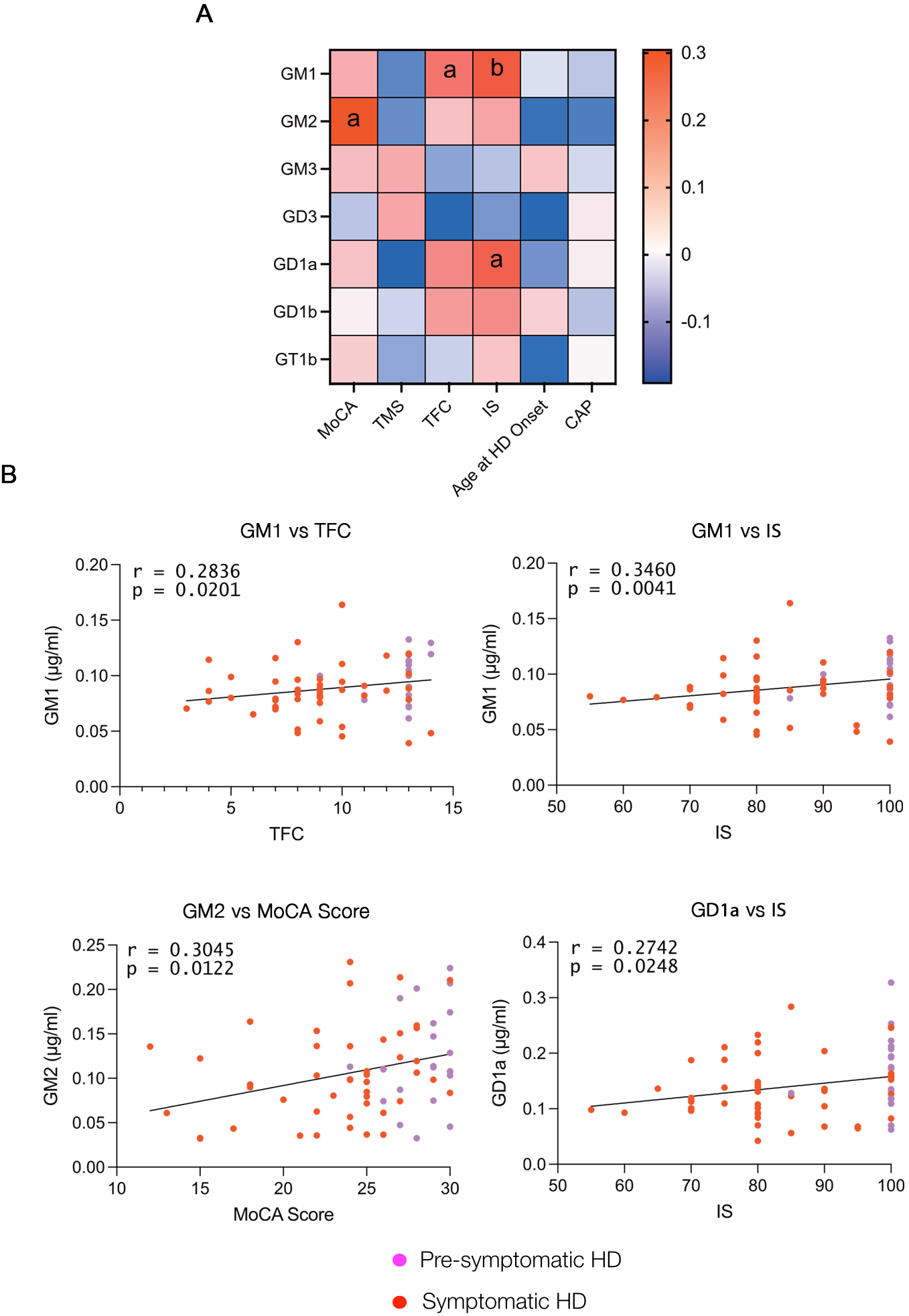
Correlation of plasma gangliosides and clinical scores in HD carriers. A. Heat map shows Pearson/Spearman r values. **a,** *p*<0.05; **b,** *p*<0.01. B. Graphs detail significant correlations. Purple and red dots: pre-symptomatic and symptomatic HD carriers, respectively.

To further explore these associations and quantify effect size, we performed univariate linear regression analysis. Consistent with the correlation analyses, the regression model confirmed that GM2 levels were positively associated with MoCA scores (β = 26.18, *p* = 0.012), while GM1 and GD1a levels were positively associated with IS (β = 138.712, *p* = 0.0311; and β = 52.585, *p* = 0.041, respectively). Although the associations between GM1 and TFC did not reach statistical significance in the regression model, the trend aligned with the correlation results. Thus, both correlation and regression analyses show an important link between higher levels of these gangliosides and less severe disease symptoms, highlighting their potential protective role in HD.

### Specific GM3 and GD3 ceramide species correlate with symptom severity and disease burden

Each ganglioside class includes a variety of species that share the same glycan structure but have ceramide backbones which differ in the length and degree of saturation of their long-chain sphingoid base and the N-acyl fatty acid. Although the biological significance of this diversity is still largely unclear, a few studies have shown that changes in ganglioside ceramide composition may occur during the differentiation of certain cells (27), ageing (28, 29) and in pathological conditions (30-32). Therefore, we investigated whether the ceramide structure of plasma gangliosides was affected in HD gene carriers. We detected 11 and 7 species for GM3 and GD3, respectively (supplementary **Table S2** and **Fig. 3**), but only one species (d36:1) for GM2 and GM1, and two (d36:1 and d38:1) for GD1a, GD1b and GT1b (supplementary **Table S2**). While the overall ceramide composition of GM3 and GD3 was similar between controls and HD gene carriers, the GM3 species d42:2 and d42:1 and GD3 d34:1 were more represented in HD gene carriers than in controls (**Fig. 3A** and **C**). Interestingly, although the levels of total GM3 did not appear to correlate with any clinical scores in our studies, we found that higher levels of GM3 d42:2h (aka d18:1-h24:1), a low abundance hydroxylated ganglioside, correlated with higher MoCA scores (r = 0.280, *p* = 0.022). On the other hand, higher levels of GM3 d34:1 (d18:1-16:0) correlated with higher TMS (r = 0.275, *p* = 0.024) (supplementary **Table S5**). Analysis of the GD3 species showed an inverse correlation between GD3 d34:1, the most abundant GD3 species, and TFC (r = -0.587, *p* = 0.038), and a positive correlation between GD3 d36:1 and CAP score (supplementary **Table S6**), suggesting an overall association between higher plasma GD3 levels and higher disease burden in HD patients.

**Figure 3.**
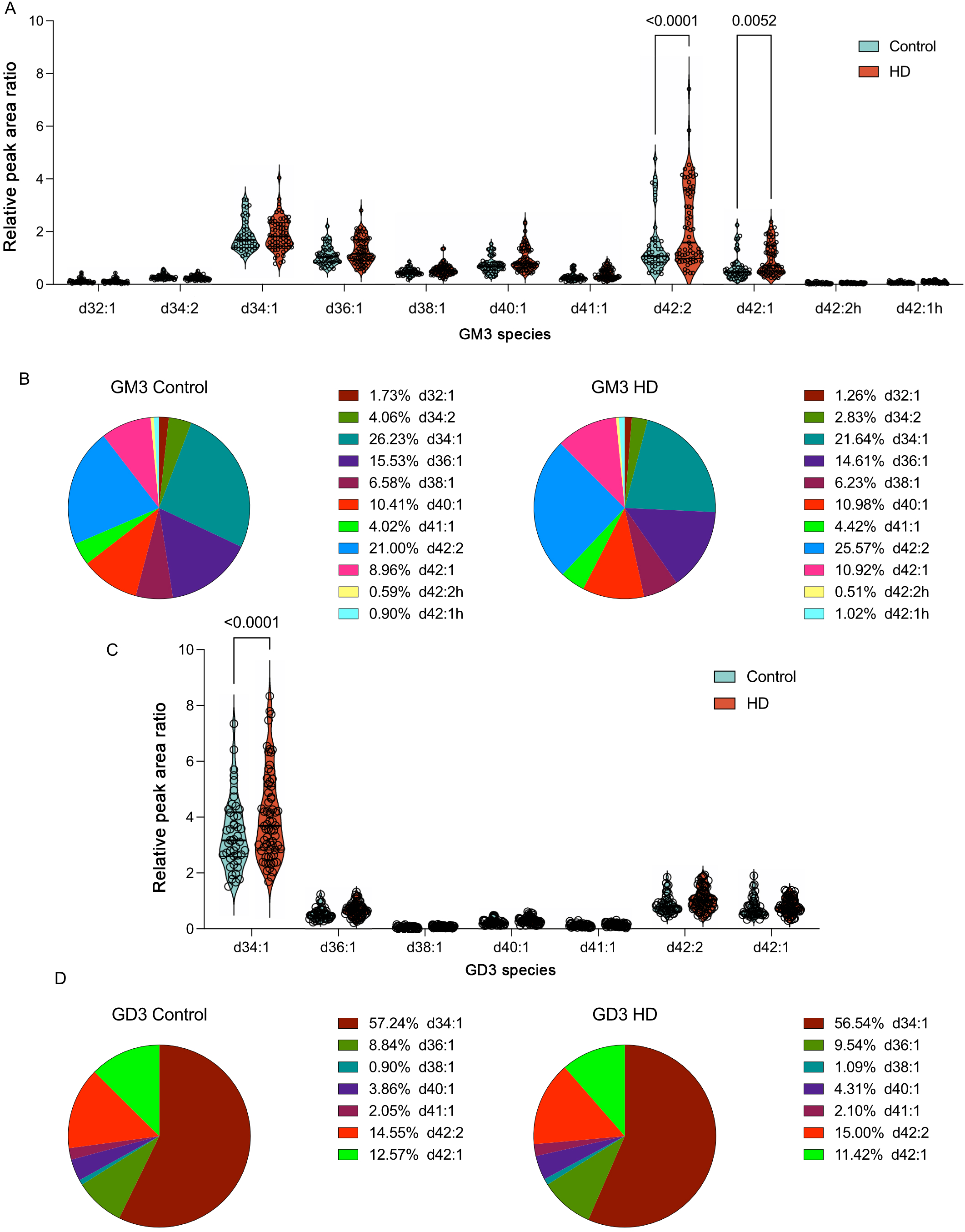
Plasma levels of GM3 and GD3 species in controls and HD carriers. Comparison of GM3 (A) and GD3 (C) species, with species distribution shown as a fraction of total GM3 (B) or GD3 (D). Relative peak area ratios were calculated by dividing the peak area of the analyte by the peak area of an internal standard. Two-way ANOVA with False Discovery Rate correction.

### Higher plasma levels of GM3 and GM1 are associated with HD status

To better understand the relationship between ganglioside levels and HD status, we initially performed univariate logistic regression for all major ganglioside classes to identify potential ganglioside biomarkers, followed by a Sure Screening variable selection method to account for potential confounding factors and intercorrelations, and identify the most predictive biomarkers in an unbiased manner. This adjusted model revealed significant associations between different gangliosides and disease risk. We found higher plasma levels of GM3 and GM1, and lower levels of GD3 to be associated with higher odds to be an HD-carrier (**Table 2**), an association that was also confirmed using a standard multivariate logistic regression model (data not shown).

**Table 2.**
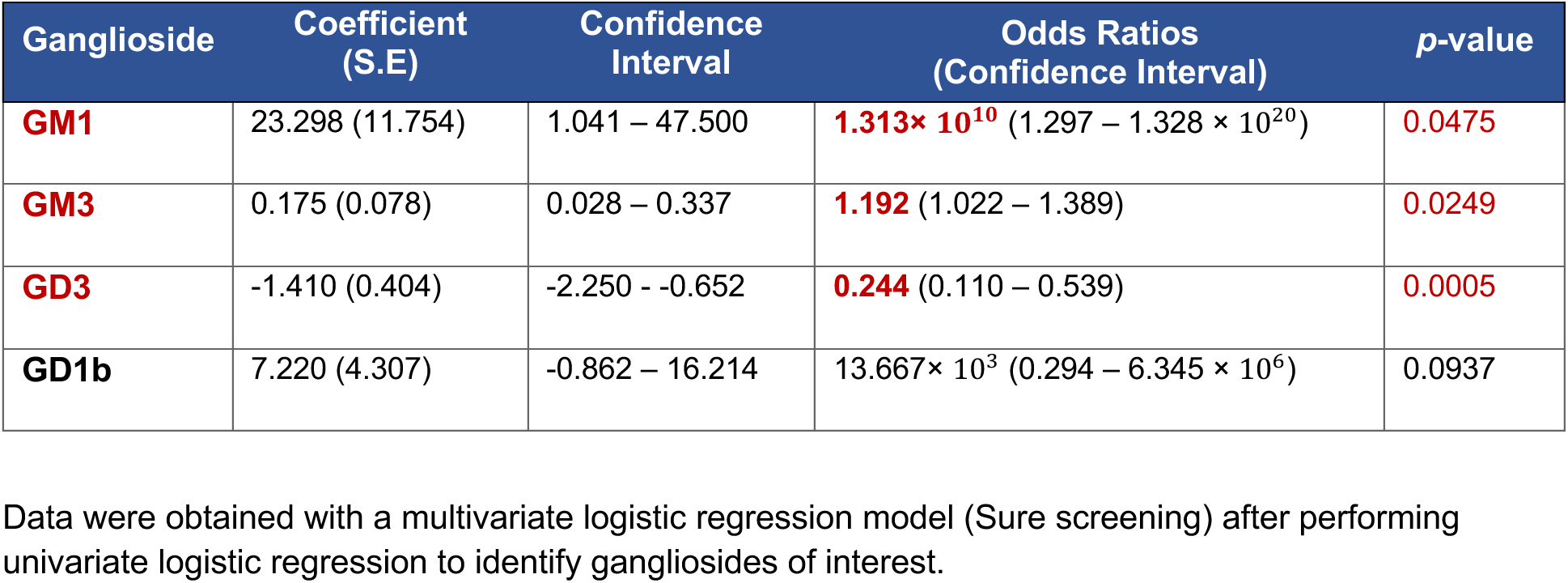
Relationship between plasma gangliosides and HD genotype.

| Ganglioside | Coefficient<br>(S.E) | Confidence<br>Interval | Odds Ratios<br>(Confidence Interval) | p-value |
| --- | --- | --- | --- | --- |
| **GM1** | 23.298 (11.754) | 1.041 – 47.500 | **1.313× 10<sup>10</sup>** (1.297 – 1.328 × 10<sup>20</sup>) | **0.0475** |
| **GM3** | 0.175 (0.078) | 0.028 – 0.337 | **1.192** (1.022 – 1.389) | **0.0249** |
| **GD3** | -1.410 (0.404) | -2.250 - -0.652 | **0.244** (0.110 – 0.539) | **0.0005** |
| **GD1b** | 7.220 (4.307) | -0.862 – 16.214 | 13.667× 10<sup>3</sup> (0.294 – 6.345 × 10<sup>6</sup>) | 0.0937 |

Data were obtained with a multivariate logistic regression model (Sure screening) after performing univariate logistic regression to identify gangliosides of interest.

### Declining GM1 levels and rising GD3 predict disease progression

Next, we investigated the relationship between plasma ganglioside levels and HD progression from early presymptomatic to late presymptomatic, mild and moderate symptomatic stages, correcting for potential effects of age and sex. Multivariate logistic regression revealed GM1 decline as a strong predictor of disease progression, while higher GD3 levels were associated with more advanced HD stages (**Table 3**).

**Table 3.** Relationship between plasma gangliosides and HD stage.

| Variable | Coefficient<br>(S.E) | Odds Ratios | Confidence Interval | p-value |
| --- | --- | --- | --- | --- |
| **Sex (Female)** | Reference |  |  |  |
| **Sex (Male)** | 0.4482 (0.527) | 1.565 | (0.5594 – 4.4779) | 0.3953 |
| **Age at visit** | **0.0882 (0.023)** | **1.092** | (1.0446 – 1.1469) | **0.0002** |
| **GM3** | -0.0157 (0.098) | 0.984 | (0.8083 – 1.1960) | 0.8731 |
| **GD3** | **1.4475 (0.604)** | **4.253** | (1.3509 – 14.2859) | **0.0166** |
| **GT1b** | 0.0722 (6.080) | 1.075 | (0.0 – 2.1253) | 0.9905 |
| **GM1** | **-60.1149 (22.486)** | **<0.0001** | (0.0 – 0.0) | **0.0075** |
| **GM2** | 3.2751 (5.882) | 26.446 | (0.0002 – 3.0604) | 0.5777 |
| **GD1a** | 4.2639 (7.590) | 71.087 | (0.0 – 2.5896) | 0.5743 |

Data were obtained with a multivariate ordinal logistic regression model for early symptomatic vs late pre-symptomatic vs mild vs moderate HD carriers, without proportional odds assumption. Independent variables were age at visit, sex, levels of GM3, GD3, GT1b, GM1, GM2 and GD1a.

Finally, we performed Receiver Operating Characteristic (ROC) analysis to evaluate the ability of plasma ganglioside levels to differentiate HD patients from controls. GM3, GD3 and GT1b had the highest diagnostic performance among all gangliosides (AUC: 0.640, 0.614, 0.621, respectively) (supplementary **Fig. S4**). GM3 had the highest sensitivity (86.6%) but also high false positive rate (56.8%). Overall, the modest AUC values indicate that gangliosides have weak discriminatory ability to distinguish HD gene carriers from controls, below the threshold typically considered clinically relevant (AUC > 0.7).

## DISCUSSION

This is the first study to analyze plasma gangliosides in HD gene carriers, and one of very few to evaluate their diagnostic and prognostic potential in neurodegenerative conditions (33-37). Although high interindividual variability limits the diagnostic potential of plasma gangliosides, our study reveals important associations between ganglioside levels and disease severity and progression, highlighting their potential prognostic value in HD.

Higher GM2, GM1 and GD1a levels correlated with better cognitive, motor and-functional scores, suggesting that individuals with higher levels of these gangliosides may be more likely to experience milder symptoms or slower disease progression. While plasma gangliosides primarily originate from the liver (38), and their levels do not directly reflect brain concentrations, our findings align with preclinical studies that showed that increasing brain ganglioside levels through administration of GM1 improves motor and cognitive symptoms in HD mouse models and provides neuroprotection (14, 15).

Importantly, our studies revealed that higher GM1 levels predict HD status, while a decline in GM1 levels is a strong predictor of disease progression. This suggests that GM1 synthesis may be upregulated as a compensatory neuroprotective response in HD gene carriers, but this response, along with GM1 levels, progressively declines in later disease stages. A similar hypothesis was proposed to explain the role of GM3 elevation in amyotrophic lateral sclerosis (ALS) (39), given that despite increased GM3 levels in the spinal cord of ALS patients and animal models, inhibition of ganglioside synthesis worsened disease progression, whereas exogenous GM3 administration was protective in ALS mice (39). Whether GM3 itself was protective or if its effects were due to its potential conversion into GM1 and other complex gangliosides remains unclear.

In our study, we also observed a statistically significant increase in plasma GM3 levels in HD carriers compared to controls, with higher GM3 levels predicting HD status. Unlike the ALS study (39), GM3 is unlikely to be protective in HD, as GM3 d34:1 correlated with motor impairment. Furthermore, our unpublished *in vitro* data indicate that GM3 does not exert neuroprotective effects in HD cell models.

An increase in plasma GM3 was also reported in idiopathic Parkinson’s disease (33), suggesting that this phenomenon may not be specific to HD, but could be a feature of neurodegenerative diseases more broadly. Higher GM3 levels have also been observed in individuals with type 2 diabetes and obesity (40). While our cohort did not include patients with overt diabetes, we did not evaluate metabolic parameters such as body weight, BMI or markers of metabolic syndrome, leaving the possibility that concomitant metabolic alterations may have contributed to plasma GM3 levels in our study.

Although total GM3 levels did not correlate with any clinical variable, individual GM3 species showed distinct associations: as mentioned above, GM3 d34:1 (∼22% of total plasma GM3 in HD carriers) correlated positively with higher TMS, while GM3 d42:2h - a low abundance hydroxylated species that was found to be elevated in individuals with metabolic syndrome (41) - correlated positively with MoCA scores. The biological significance of these correlations remains unclear, as little is known on the impact of ceramide variation on ganglioside physiology.

In parallel with a GM1 decline, higher plasma GD3 levels also predicted HD progression in our study, with an increase in GD3 d34:1 and d36:1 correlating with worse motor function and increased disease burden and progression risk (as measured by the CAP score), respectively. GD3 is known to exert pro-apoptotic activity in certain cell contexts (42-45), but whether this might contribute to HD progression is not known.

Overall, while plasma gangliosides are weak disease classifiers and insufficient as standalone biomarkers for HD diagnosis, their associations with disease severity and progression suggest that they play a role in HD pathogenesis and may hold prognostic value. Our regression models revealed a strong association between declining GM1 levels and disease progression. Future longitudinal studies could further clarify this relationship by determining whether the rate of ganglioside depletion correlates with the rate of disease progression. Moreover, analysis of gangliosides in the cerebrospinal fluid could provide a more direct and sensitive measure of brain ganglioside changes occurring during the course of the disease.

In conclusion, our study provides important insights into ganglioside involvement in HD, corroborating preclinical data and supporting the idea that increasing GM1 levels in HD patients may offer therapeutic benefits. Our findings might have broader relevance, given the potential role of gangliosides in other conditions, with Parkinson’s disease being a prominent example.

## Data Availability

All data produced in the present study are available upon reasonable request to the authors

## ACKNOWLEDGMENTS

We are grateful to all study participants for their time and contributions to this research. We thank Dr. Sylvain Chouinard (University of Montreal) for providing a subset of plasma samples and clinical data used in this study, and the Alberta SPOR SUPPORT Unit and EPICORE at the University of Alberta for statistical support services. This work was supported by a Brain Canada Multi-Investigator Research Initiative grant to SS, OS, SC, JC and JS, and by Canadian Institutes of Health Research (CIHR) and GlycoNET grants to SS.

## AUTHORS’ ROLES

SS and OS designed study; OS and JS performed neurological tests and collected samples; DG, ZH, BP and BV performed analysis; SS, JC and YAH supervised experiments and/or analysis, SS, DG and AZ wrote the manuscript; all authors read and approved the final version of the manuscript.

## REFERENCES

1. Sipione S, Monyror J, Galleguillos D, Steinberg N, Kadam V. Gangliosides in the Brain: Physiology, Pathophysiology and Therapeutic Applications. Front Neurosci. 2020;14:572965.

2. Simpson MA, Cross H, Proukakis C, Priestman DA, Neville DC, Reinkensmeier G, et al. Infantile-onset symptomatic epilepsy syndrome caused by a homozygous loss-of-function mutation of GM3 synthase. Nat Genet. 2004;36(11):1225–9.

3. Boukhris A, Schule R, Loureiro JL, Lourenco CM, Mundwiller E, Gonzalez MA, et al. Alteration of ganglioside biosynthesis responsible for complex hereditary spastic paraplegia. Am J Hum Genet. 2013;93(1):118–23.

4. Harlalka GV, Lehman A, Chioza B, Baple EL, Maroofian R, Cross H, et al. Mutations in B4GALNT1 (GM2 synthase) underlie a new disorder of ganglioside biosynthesis. Brain. 2013;136(Pt 12):3618–24.

5. Kracun I, Rosner H, Drnovsek V, Vukelic Z, Cosovic C, Trbojevic-Cepe M, Kubat M. Gangliosides in the human brain development and aging. Neurochem Int. 1992;20(3):421–31.

6. Mo L, Ren Q, Duchemin AM, Neff NH, Hadjiconstantinou M. GM1 and ERK signaling in the aged brain. Brain Res. 2005;1054(2):125–34.

7. Palestini P, Masserini M, Sonnino S, Giuliani A, Tettamanti G. Changes in the ceramide composition of rat forebrain gangliosides with age. J Neurochem. 1990;54(1):230–5.

8. Segler-Stahl K, Webster JC, Brunngraber EG. Changes in the concentration and composition of human brain gangliosides with aging. Gerontology. 1983;29(3):161–8.

9. Wu G, Lu ZH, Kulkarni N, Ledeen RW. Deficiency of ganglioside GM1 correlates with Parkinson’s disease in mice and humans. J Neurosci Res. 2012;90(10):1997–2008.

10. Maglione V, Marchi P, Di Pardo A, Lingrell S, Horkey M, Tidmarsh E, Sipione S. Impaired ganglioside metabolism in Huntington’s disease and neuroprotective role of GM1. J Neurosci. 2010;30(11):4072–80.

11. Desplats PA, Denny CA, Kass KE, Gilmartin T, Head SR, Sutcliffe JG, et al. Glycolipid and ganglioside metabolism imbalances in Huntington’s disease. Neurobiol Dis. 2007;27(3):265–77.

12. Blennow K, Davidsson P, Wallin A, Fredman P, Gottfries CG, Karlsson I, et al. Gangliosides in cerebrospinal fluid in ‘probable Alzheimer’s disease’. Arch Neurol. 1991;48(10):1032–5.

13. Blennow K, Davidsson P, Wallin A, Fredman P, Gottfries CG, Mansson JE, Svennerholm L. Differences in cerebrospinal fluid gangliosides between “probable Alzheimer’s disease” and normal aging. Aging (Milano). 1992;4(4):301–6.

14. Di Pardo A, Maglione V, Alpaugh M, Horkey M, Atwal RS, Sassone J, et al. Ganglioside GM1 induces phosphorylation of mutant huntingtin and restores normal motor behavior in Huntington disease mice. Proc Natl Acad Sci U S A. 2012;109(9):3528–33.

15. Alpaugh M, Galleguillos D, Forero J, Morales LC, Lackey SW, Kar P, et al. Disease-modifying effects of ganglioside GM1 in Huntington’s disease models. EMBO Mol Med. 2017;9(11):1537–57.

16. Tabrizi SJ, Reilmann R, Roos RA, Durr A, Leavitt B, Owen G, et al. Potential endpoints for clinical trials in premanifest and early Huntington’s disease in the TRACK-HD study: analysis of 24 month observational data. Lancet Neurol. 2012;11(1):42–53.

17. Tabrizi SJ, Scahill RI, Owen G, Durr A, Leavitt BR, Roos RA, et al. Predictors of phenotypic progression and disease onset in premanifest and early-stage Huntington’s disease in the TRACK-HD study: analysis of 36-month observational data. Lancet Neurol. 2013;12(7):637–49.

18. Pan W, Wang X, Xiao W, Zhu H. A Generic Sure Independence Screening Procedure. J Am Stat Assoc. 2019;114(526):928–37.

19. Fan J, Lv J. Sure independence screening for ultrahigh dimensional feature space. Journal of the Royal Statistical Society: Series B (Statistical Methodology). 2008;70(5):849–911.

20. Saldana DF, Feng Y. SIS: An R Package for Sure Independence Screening in Ultrahigh-Dimensional Statistical Models. Journal of Statistical Software. 2018;83(2):1–25.

21. Unified Huntington’s Disease Rating Scale: reliability and consistency. Huntington Study Group. Mov Disord. 1996;11(2):136-42.

22. Penney Jr JB, Vonsattel J-P, Macdonald ME, Gusella JF, Myers RH. CAG repeat number governs the development rate of pathology in Huntington’s disease. Annals of Neurology. 1997;41(5):689–92.

23. Rosca EC, Simu M. Montreal cognitive assessment for evaluating cognitive impairment in Huntington’s disease: a systematic review. CNS Spectr. 2022;27(1):27–45.

24. Huang Q, Zhou X, Liu D, Xin B, Cechner K, Wang H, Zhou A. A new liquid chromatography/tandem mass spectrometry method for quantification of gangliosides in human plasma. Analytical Biochemistry. 2014;455:26–34.

25. Sibille E, Berdeaux O, Martine L, Bron AM, Creuzot-Garcher CP, He Z, et al. Ganglioside Profiling of the Human Retina: Comparison with Other Ocular Structures, Brain and Plasma Reveals Tissue Specificities. PLoS One. 2016;11(12):e0168794.

26. Li Q, Sun M, Yu M, Fu Q, Jiang H, Yu G, Li G. Gangliosides profiling in serum of breast cancer patient: GM3 as a potential diagnostic biomarker. Glycoconj J. 2019;36(5):419–28.

27. Go S, Go S, Veillon L, Ciampa MG, Mauri L, Sato C, et al. Altered expression of ganglioside GM3 molecular species and a potential regulatory role during myoblast differentiation. J Biol Chem. 2017;292(17):7040–51.

28. Caughlin S, Maheshwari S, Weishaupt N, Yeung KK, Cechetto DF, Whitehead SN. Age-dependent and regional heterogeneity in the long-chain base of A-series gangliosides observed in the rat brain using MALDI Imaging. Sci Rep. 2017;7(1):16135.

29. Sugiura Y, Shimma S, Konishi Y, Yamada MK, Setou M. Imaging mass spectrometry technology and application on ganglioside study; visualization of age-dependent accumulation of C20-ganglioside molecular species in the mouse hippocampus. PLoS One. 2008;3(9):e3232.

30. Caughlin S, Maheshwari S, Agca Y, Agca C, Harris AJ, Jurcic K, et al. Membrane-lipid homeostasis in a prodromal rat model of Alzheimer’s disease: Characteristic profiles in ganglioside distributions during aging detected using MALDI imaging mass spectrometry. Biochim Biophys Acta Gen Subj. 2018;1862(6):1327–38.

31. Hirano-Sakamaki W, Sugiyama E, Hayasaka T, Ravid R, Setou M, Taki T. Alzheimer’s disease is associated with disordered localization of ganglioside GM1 molecular species in the human dentate gyrus. FEBS Lett. 2015;589(23):3611–6.

32. Kanoh H, Nitta T, Go S, Inamori KI, Veillon L, Nihei W, et al. Homeostatic and pathogenic roles of GM3 ganglioside molecular species in TLR4 signaling in obesity. Embo j. 2020;39(12):e101732.

33. Chan RB, Perotte AJ, Zhou B, Liong C, Shorr EJ, Marder KS, et al. Elevated GM3 plasma concentration in idiopathic Parkinson’s disease: A lipidomic analysis. PLoS One. 2017;12(2):e0172348.

34. Alselehdar SK, Chakraborty M, Chowdhury S, Alcalay RN, Surface M, Ledeen R. Subnormal GM1 in PBMCs: Promise for Early Diagnosis of Parkinson’s Disease? Int J Mol Sci. 2021;22(21).

35. Avisar H, Guardia-Laguarta C, Area-Gomez E, Surface M, Chan AK, Alcalay RN, Lerner B. Lipidomics Prediction of Parkinson’s Disease Severity: A Machine-Learning Analysis. J Parkinsons Dis. 2021;11(3):1141–55.

36. Ledeen R, Chowdhury S, Lu ZH, Chakraborty M, Wu G. Systemic deficiency of GM1 ganglioside in Parkinson’s disease tissues and its relation to the disease etiology. Glycoconj J. 2022;39(1):75–82.

37. Zhang J, Zhang X, Wang L, Yang C. High Performance Liquid Chromatography-Mass Spectrometry (LC-MS) Based Quantitative Lipidomics Study of Ganglioside-NANA-3 Plasma to Establish Its Association with Parkinson’s Disease Patients. Med Sci Monit. 2017;23:5345–53.

38. Jennemann R, Rothermel U, Wang S, Sandhoff R, Kaden S, Out R, et al. Hepatic Glycosphingolipid Deficiency and Liver Function in Mice. Hepatology. 2010;51(5):1799–809.

39. Dodge JC, Treleaven CM, Pacheco J, Cooper S, Bao C, Abraham M, et al. Glycosphingolipids are modulators of disease pathogenesis in amyotrophic lateral sclerosis. Proc Natl Acad Sci U S A. 2015;112(26):8100–5.

40. Sato T, Nihei Y, Nagafuku M, Tagami S, Chin R, Kawamura M, et al. Circulating levels of ganglioside GM3 in metabolic syndrome: A pilot study. Obesity Research & Clinical Practice. 2008;2(4):231–8.

41. Veillon L, Go S, Matsuyama W, Suzuki A, Nagasaki M, Yatomi Y, Inokuchi J. Identification of Ganglioside GM3 Molecular Species in Human Serum Associated with Risk Factors of Metabolic Syndrome. PLoS One. 2015;10(6):e0129645.

42. Garofalo T, Giammarioli AM, Misasi R, Tinari A, Manganelli V, Gambardella L, et al. Lipid microdomains contribute to apoptosis-associated modifications of mitochondria in T cells. Cell Death Differ. 2005;12(11):1378–89.

43. Sorice M, Mattei V, Matarrese P, Garofalo T, Tinari A, Gambardella L, et al. Dynamics of mitochondrial raft-like microdomains in cell life and death. Commun Integr Biol. 2012;5(2):217–9.

44. Simon BM, Malisan F, Testi R, Nicotera P, Leist M. Disialoganglioside GD3 is released by microglia and induces oligodendrocyte apoptosis. Cell Death Differ. 2002;9(7):758–67.

45. Melchiorri D, Martini F, Lococo E, Gradini R, Barletta E, De Maria R, et al. An early increase in the disialoganglioside GD3 contributes to the development of neuronal apoptosis in culture. Cell Death Differ. 2002;9(6):609–15.

