## Supplementary Materials for "Plasma gangliosides correlate with disease stages and symptom severity in Huntington’s disease carriers"

#### SUPPLEMENTARY FIGURE LEGENDS

**Fig. S1 - Plasma ganglioside levels do not correlate with study participants' age.** The association between the levels of gangliosides in plasma and participants' age was calculated using Pearson's correlation. Ganglioside plasma levels of each participant are presented in grey circles for healthy controls, purple for pre-symptomatic individuals and red for symptomatic patients.

**Fig S2 - Plasma ganglioside levels do not significantly differ between male and female participants.** Box plots show the median, interquartile range, minimum and maximum values (whiskers) of plasma ganglioside levels in male and female participants. Numbers are *p*-values. Mann-Whitney U test, with a statistical significance threshold set at $p<0.05$.

**Fig S3 - Plasma ganglioside levels do not correlate with CAG repeat length in HD carriers.** The correlation coefficient between the levels of gangliosides in plasma and participants' CAG length was calculated using Pearson's correlation. Ganglioside plasma levels of each participant are presented in grey circles for healthy controls, purple for pre-symptomatic individuals and red for symptomatic patients.

**Fig S4 – ROC Analysis.** Receiver operating characteristic (ROC) analysis for the major plasma gangliosides. The table shows the summary of results. ROC curves for the most relevant gangliosides (GM1 and those with the highest AUC) are shown at the bottom. The optimal Youden Index identifies the threshold that provides the best balance between sensitivity and specificity. AUC, area under the curve; CI, confidence interval; TPR, true positive rate; FPR, false positive rate.

### Figure S1

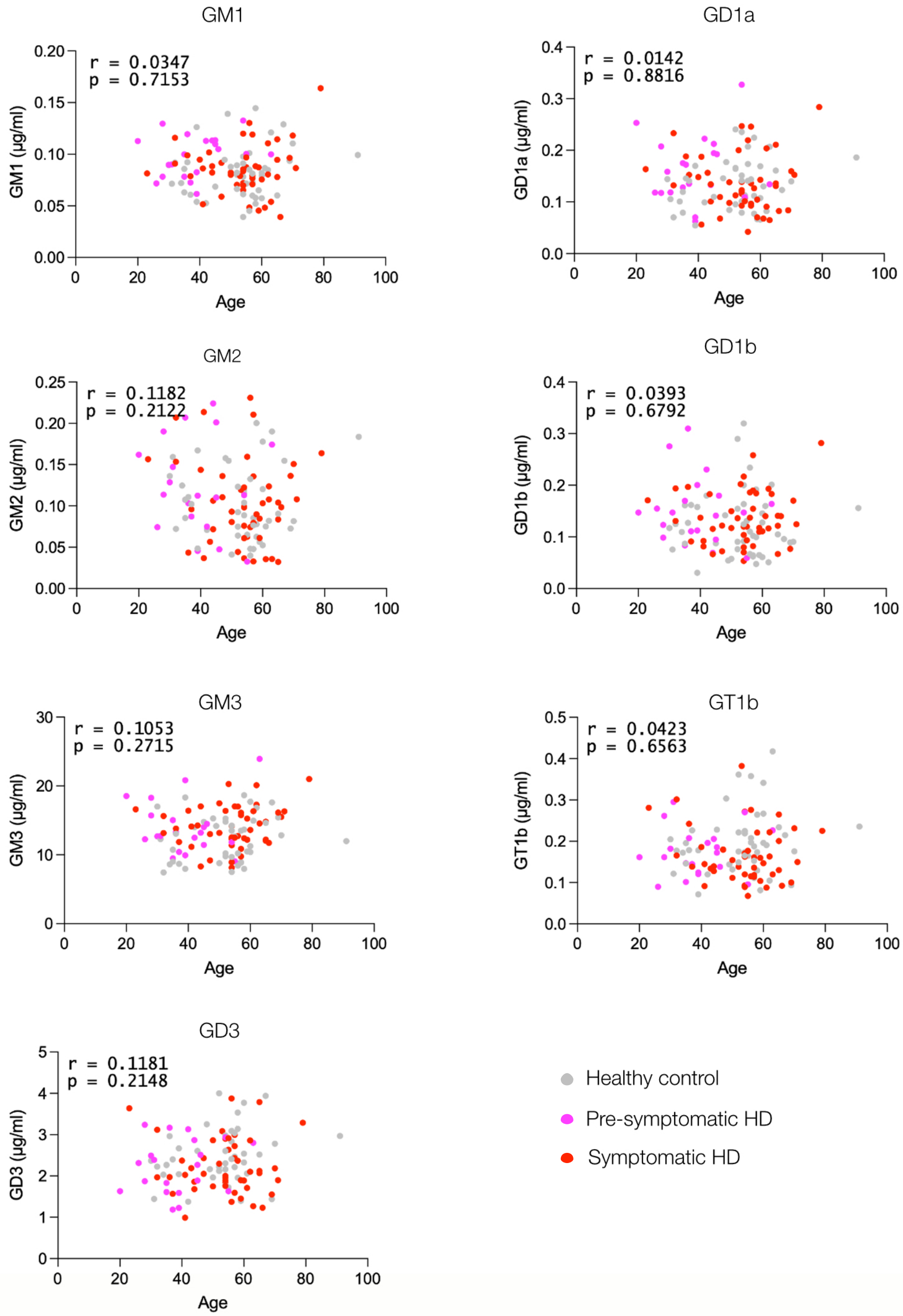
GM1
r = 0.0347
p = 0.7153
GM1 (µg/ml)
Age
GD1a
r = 0.0142
p = 0.8816
GD1a (µg/ml)
Age
GM2
r = 0.1182
p = 0.2122
GM2 (µg/ml)
Age
GD1b
r = 0.0393
p = 0.6792
GD1b (µg/ml)
Age
GM3
r = 0.1053
p = 0.2715
GM3 (µg/ml)
Age
GT1b
r = 0.0423
p = 0.6563
GT1b (µg/ml)
Age
GD3
r = 0.1181
p = 0.2148
GD3 (µg/ml)
Age
Healthy control
Pre-symptomatic HD
Symptomatic HD

#### Figure S2

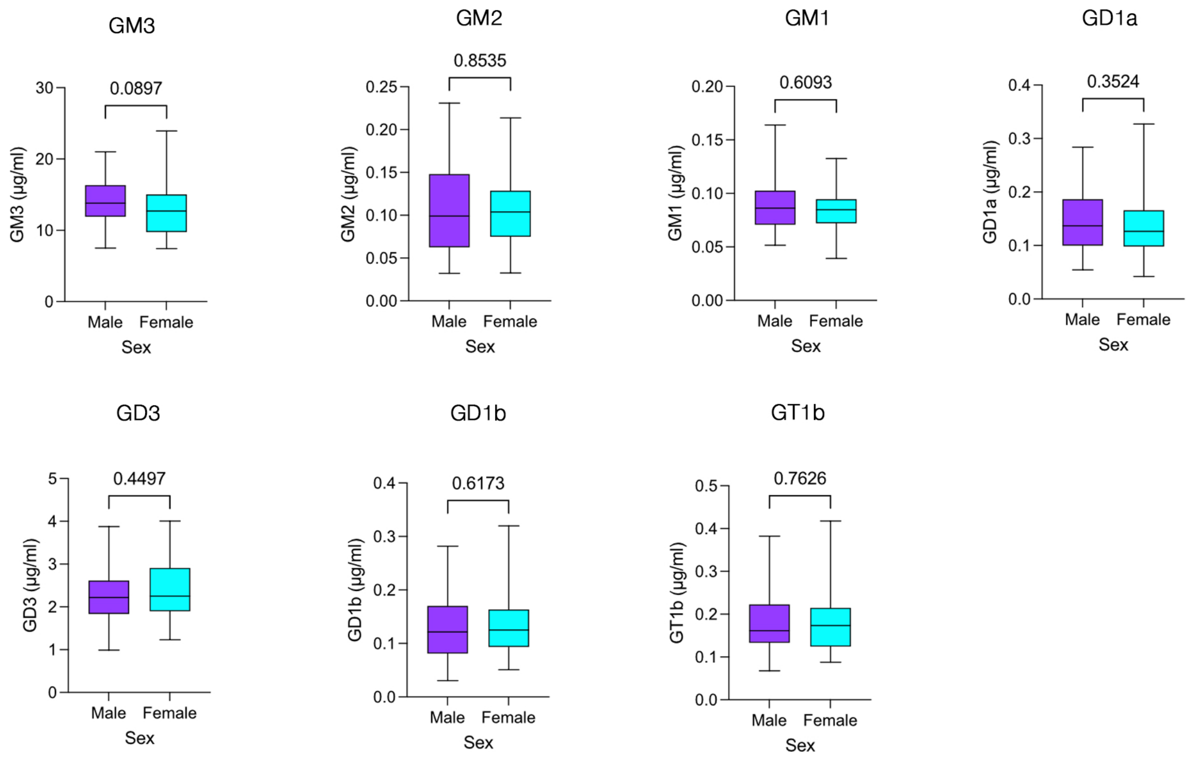
GM3
0.0897
GM3 (µg/ml)
30
20
10
0
Male
Female
Sex
GM2
0.8535
GM2 (µg/ml)
0.25
0.20
0.15
0.10
0.05
0.00
Male
Female
Sex
GM1
0.6093
GM1 (µg/ml)
0.20
0.15
0.10
0.05
0.00
Male
Female
Sex
GD1a
0.3524
GD1a (µg/ml)
0.4
0.3
0.2
0.1
0.0
Male
Female
Sex
GD3
0.4497
GD3 (µg/ml)
5
4
3
2
1
0
Male
Female
Sex
GD1b
0.6173
GD1b (µg/ml)
0.4
0.3
0.2
0.1
0.0
Male
Female
Sex
GT1b
0.7626
GT1b (µg/ml)
0.5
0.4
0.3
0.2
0.1
0.0
Male
Female
Sex

#### Figure S3

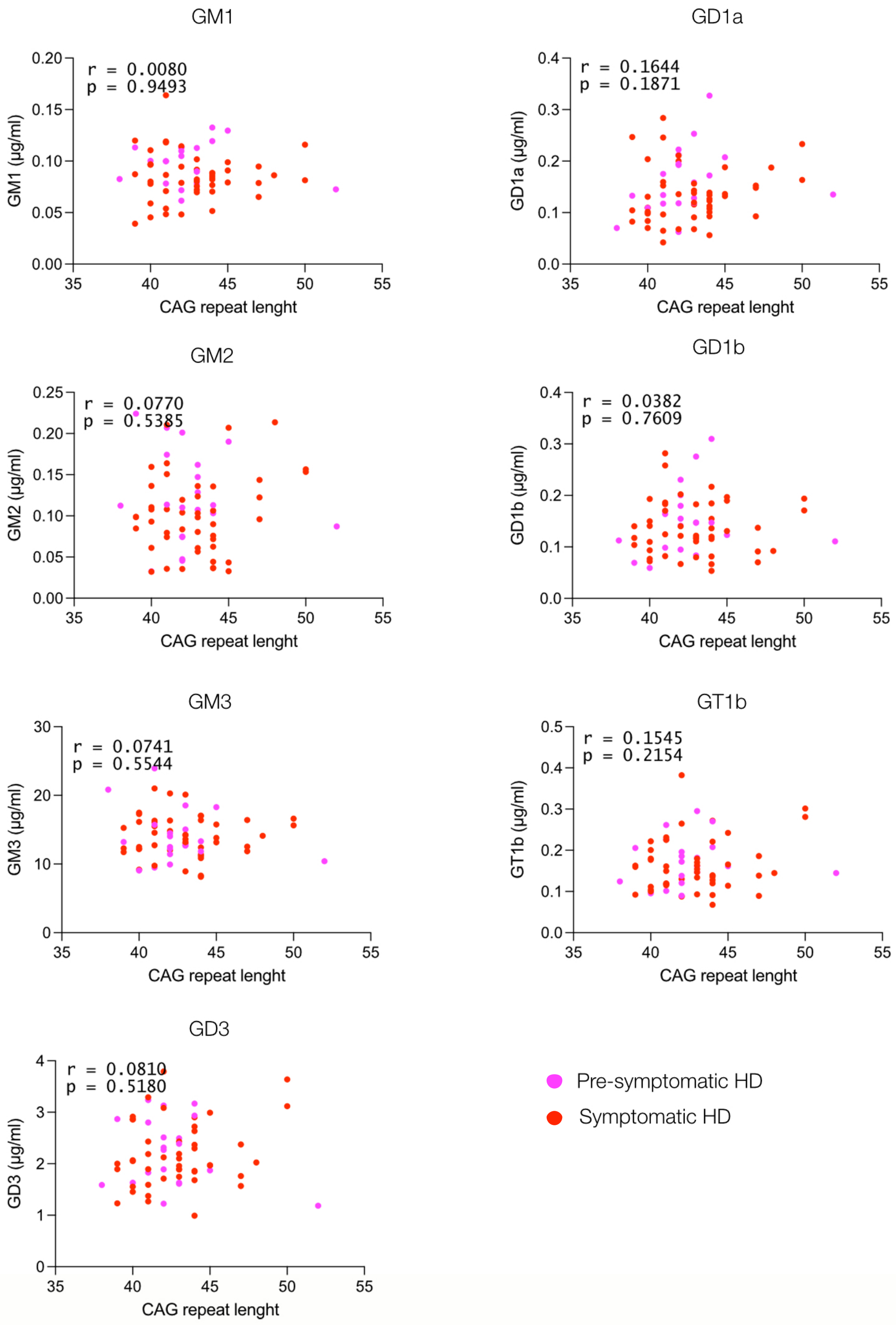

GM1
r = 0.0080
p = 0.9493
GM1 (µg/ml)
CAG repeat lenght
GD1a
r = 0.1644
p = 0.1871
GD1a (µg/ml)
CAG repeat lenght
GM2
r = 0.0770
p = 0.5385
GM2 (µg/ml)
CAG repeat lenght
GD1b
r = 0.0382
p = 0.7609
GD1b (µg/ml)
CAG repeat lenght
GM3
r = 0.0741
p = 0.5544
GM3 (µg/ml)
CAG repeat lenght
GT1b
r = 0.1545
p = 0.2154
GT1b (µg/ml)
CAG repeat lenght
GD3
r = 0.0810
p = 0.5180
GD3 (µg/ml)
CAG repeat lenght
Pre-symptomatic HD
Symptomatic HD

### Figure S4

| Ganglioside | Optimal Youden Index | 1-Specificity (FPR) | Sensitivity (TPR) | AUC (95% CI) | Interpretation |
|---|---|---|---|---|---|
| GM1 | 0.1924 | 0.1957 | 0.3881 | 0.599 (0.491 - 0.707) | Poor discrimination |
| GM2 | 0.1337 | 0.4783 | 0.6119 | 0.525 (0.417 - 0.633) | Poor discrimination |
| GM3 | 0.2975 | 0.5682 | 0.8657 | 0.640 (0.531 - 0.749) | Weak-moderate discrimination |
| GD3 | 0.2998 | 0.5224 | 0.8222 | 0.615 (0.509 – 0.719) | Weak-moderate discrimination |
| GD1a | 0.1450 | 0.1087 | 0.2537 | 0.548 (0.440 - 0.657) | Poor discrimination |
| GD1b | 0.2382 | 0.4783 | 0.7164 | 0.606 (0.497 - 0.716) | Weak discrimination |
| GT1b | 0.3008 | 0.3731 | 0.6739 | 0.621 (0.515 - 0.726) | Weak-moderate discrimination |

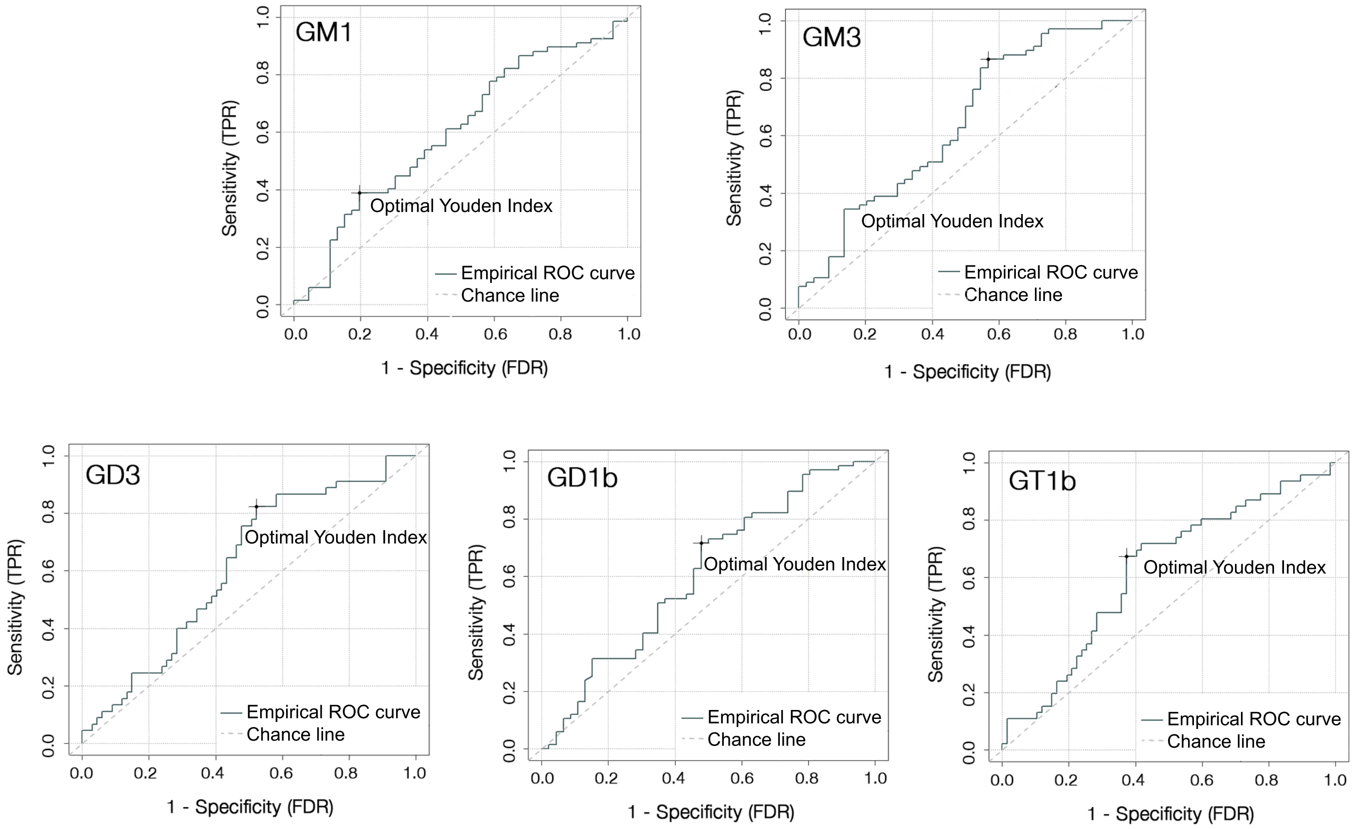

**Table S1. Summary of MS-MS experimental conditions and ganglioside species detected**

| Class | MRM | Species | Molecular | Formula | DP | EP | CE | CXP | CEP |
|---|---|---|---|---|---|---|---|---|---|
| GM1, [M-H]- | 1544.8/290 | d36:1 | d18:1-18:0 | $C_{73}H_{131}N_3O_{31}$ | -165 | -10 | -105 | -2 | -69 |
| GM2, [M-H]- | 1382.8/290 | d36:1 | d18:1-18:0 | $C_{67}H_{121}N_3O_{26}$ | -170 | -10 | -85 | -2 | -63 |
| GM3, [M-H]- | 1123.6/290 | d32:1 | d18:1-14:0 | $C_{55}H_{100}N_2O_{21}$ | -100 | -10 | -70 | -2 | -53 |
| | 1149.6/290 | d34:2 | d18:1-16:1 | $C_{57}H_{102}N_2O_{21}$ | -100 | -10 | -70 | -2 | -54 |
| | 1151.6/290 | d34:1 | d18:1-16:0 | $C_{57}H_{104}N_2O_{21}$ | -100 | -10 | -66 | -2 | -40 |
| | 1179.8/290 | d36:1 | d18:1-18:0 | $C_{59}H_{108}N_2O_{21}$ | -100 | -10 | -70 | -2 | -55 |
| | 1207.8/290 | d38:1 | d18:1-20:0 | $C_{61}H_{112}N_2O_{21}$ | -100 | -10 | -70 | -2 | -56 |
| | 1235.8/290 | d40:1 | d18:1-22:0 | $C_{63}H_{116}N_2O_{21}$ | -85 | -12 | -72 | -2 | -40 |
| | 1249.8/290 | d41:1 | d18:1-23:0 | $C_{64}H_{118}N_2O_{21}$ | -115 | -11 | -70 | -2 | -40 |
| | 1261.7/290 | d42:2 | d18:1-24:1 | $C_{65}H_{118}N_2O_{21}$ | -100 | -10 | -70 | -2 | -58 |
| | 1263.7/290 | d42:1 | d18:1-24:0 | $C_{65}H_{120}N_2O_{21}$ | -100 | -10 | -70 | -2 | -58 |
| | 1277.8/290 | d42:2h | d18:1-h24:1 | $C_{66}H_{122}N_2O_{21}$ | -100 | -10 | -70 | -2 | -59 |
| | 1279.8/290 | d42:1h | d18:1-h24:0 | $C_{66}H_{124}N_2O_{21}$ | -100 | -10 | -70 | -2 | -59 |
| GD3, [M-2H]2- | 720.8/290.1 | d34:1 | d18:1-16:0 | $C_{68}H_{121}N_3O_{29}$ | -60 | -10 | -46 | -2 | -38 |
| | 734.9/290.1 | d36:1 | d18:1-18:0 | $C_{70}H_{125}N_3O_{29}$ | -60 | -10 | -46 | -2 | -39 |
| | 748.9/290.1 | d38:1 | d18:1-20:0 | $C_{72}H_{129}N_3O_{29}$ | -80 | -8 | -50 | -2 | -30 |
| | 762.9/290.1 | d40:1 | d18:1-22:0 | $C_{74}H_{133}N_3O_{29}$ | -60 | -10 | -46 | -2 | -30 |
| | 769.9/290.1 | d41:1 | d18:1-23:0 | $C_{75}H_{135}N_3O_{29}$ | -70 | -10 | -50 | -2 | -30 |
| | 775.9/290.1 | d42:2 | d18:1-24:1 | $C_{76}H_{135}N_3O_{29}$ | -60 | -10 | -46 | -2 | -40 |
| | 776.9/290.1 | d42:1 | d18:1-24:0 | $C_{76}H_{137}N_3O_{29}$ | -70 | -10 | -45 | -2 | -30 |
| GD1a/b,$[M-2H]^{2-}$ | 917.4/290.1 | d36:1 | d18:1-18:0 | $C_{84}H_{148}N_4O_{39}$ | -95 | -10 | -54 | -4 | -28 |
| | 931.5/290.1 | d38:1 | d18:1-20:0 | $C_{86}H_{152}N_4O_{39}$ | -90 | -10 | -54 | -2 | -34 |
| GT1b,$[M-2H]^{2-}$ | 1063.0/290 | d36:1 | d18:1-18:0 | $C_{95}H_{165}N_5O_{47}$ | -70 | -10 | -44 | -2 | -36 |
| | 1077.0/290 | d38:1 | d18:1-20:0 | $C_{97}H_{169}N_5O_{47}$ | -100 | -10 | -62 | -2 | -51 |

Gangliosides (Class), MRM transition ion pairs (MRM), ceramide composition (Species), molecular structures of ceramide species (Molecular), formula and mass spectrometric parameters for quantification (declustering potential, DP; entrance potential, EP; collision cell entrance potential, CE; collision energy, CXP and collision cell exit potential, CEP). d-values are the sum of the carbon atoms in the sphingoid base and the fatty acid moiety and the number of double bonds present in each detected ganglioside species.

**Table S2. Linearity and precision of HILIC-MS plasma ganglioside analysis.**

| Gangliosides | Range of linearity | Linear regression equation in pure solvent | $r^2$ | Linear regression equation in plasma matrix | $r^2$ | Matrix effect |
|---|---|---|---|---|---|---|
| GM3 | 5-500ng | y=2.02x+0.3934 | 0.9997 | y=1.35x+17.09 | 0.9888 | 66.8% |
| GM1 | 0.075-7.5 ng | y=5.57x+0.0261 | 0.9991 | y=4.13x+0.29 | 0.9995 | 74.1% |
| GD3 | 0.72-72 ng | y=3.82x+0.03 | 0.9991 | y=2.16x+3.69 | 0.9997 | 56.5% |
| GD1a | 0.92-92 ng | y=10.78x-0.046 | 0.9997 | y=15.94x+1.22 | 0.9995 | 147.8% |
| GD1b | 0.5-50 ng | y=1.01x-0.005 | 0.9996 | y=0.79x+0.10 | 0.9991 | 78.2% |
| GT1b | 0.5-50 ng | y=1.97x-0.004 | 0.9986 | y=1.9x+0.40 | 0.9999 | 96.4% |

A matrix-adjusted calibration method was developed using pooled plasma samples spiked with deuterated ganglioside standards. Pooled plasma was extracted by liquid-liquid extraction and spiked with the standard calibration solution at 5 different concentrations (plasma matrix). In parallel, a calibration curve with the same 5 standards was prepared in pure solvent. The matrix effect was obtained by dividing the calibration curve slope in plasma matrix by the slope in pure solvent.

**Table S3. Ganglioside recovery after extraction from pooled plasma samples**

| | GANGLIOSIDE RECOVERY | | | | | | |
|---|---|---|---|---|---|---|---|
| Concentration of spiked internal standards | GM3 | GM2 | GM1 | GD3 | GD1a | GD1b | GT1b |
| Low | 109% | 96% | 98% | 109% | 76% | 99% | 93% |
| High | 92% | 108% | 114% | 114% | 106% | 105% | 110% |

Deuterated internal standards were added to plasma before sample extraction with methanol. The spiked ganglioside concentrations were 0.05 µg/ml (low) and 0.5 µg/ml (high) for GM2, GM1, GD1b and GT1b; 0.025 and 0.25 µM for GD3 and GD1a; 0.2 and 2 µg/ml for GM3.
